# Serum Cotinine and Wrist-Worn Ambient Light Exposure Patterns in U.S. Adults: A Cross-Sectional Analysis of NHANES 2011–2014

**DOI:** 10.64898/2026.06.02.26354759

**Authors:** Andrew C. Wong, Cooper Lee, Adam Park, Luke Yin, Yoo Choi

**Affiliations:** University of Michigan, Ann Arbor, MI; Illinois Mathematics and Science Academy, Aurora, IL; Princeton University, Princeton, NJ; Cornell Tech, New York, NY

**Keywords:** serum cotinine, ambient light, circadian, sleep, NHANES, tobacco smoke

## Abstract

**Background:** Tobacco smoke exposure, quantified by serum cotinine, is associated with cardiovascular, metabolic, and sleep-related health risks[1, 2]. The relationship between biomarker-verified tobacco smoke exposure and objectively measured, free-living wrist-worn ambient light patterns has not been examined in a nationally representative U.S. adult sample.

**Methods:** We analyzed NHANES 2011–2014 cross-sectional data from 6,937 adults aged ≥20 years with valid serum cotinine and wrist-worn Physical Activity Monitor (PAM) ambient light data. Seven light outcomes were modeled using survey-weighted linear regression with log_2_(cotinine + 1) as the continuous exposure across four covariate adjustment levels. Benjamini–Hochberg false discovery rate (FDR) correction was applied across the 7 outcomes within each model.

**Results:** In Model 2 (adjusted for age, sex, race/ethnicity, education, poverty-income ratio, BMI, and survey cycle; *N* = 6,350), higher serum cotinine was associated with significantly higher nighttime light (*β* = +0.024, 95% CI: 0.010, 0.038; *p*_FDR_ = 0.014) and lower evening light (*β* = −0.031, 95% CI: −0.055, −0.008; *p*_FDR_ = 0.042). In exploratory behavioral models without alcohol (Model 3a; *N* = 5,766), both nighttime and evening associations remained FDR-significant. After additional adjustment for alcohol, which substantially reduced the sample due to 37.6% missingness (Model 3b; *N* = 3,866), the nighttime association attenuated below the FDR threshold, while the evening association remained FDR-significant. Categorical analyses showed progressively higher nighttime light across cotinine groups, and a hypothesis-generating sex interaction was identified (*p*_interaction_ = 0.001).

**Conclusions:** Higher serum cotinine concentrations were associated with higher nighttime and lower evening ambient light after sociodemographic adjustment. Attenuation after behavioral adjustment and the cross-sectional design preclude causal inference. Longitudinal studies with formal mediation analyses are needed to clarify the temporal ordering and mechanisms linking tobacco smoke exposure, smoking-related behaviors, and personal light–dark cycle patterns.

## 1. Introduction

Tobacco smoke is among the most consequential preventable exposures globally, contributing to excess mortality from cardiovascular disease, cancer, and respiratory conditions[1, 2]. Serum cotinine, the primary metabolite of nicotine, provides an objective biomarker of tobacco smoke exposure that captures both active smoking and passive smoke inhalation[3, 4]. Nationally representative survey data indicate that a substantial proportion of self-reported non-smokers carry detectable cotinine concentrations, reflecting widespread passive exposure[5].

Circadian disruption is recognized as an independent risk factor for cardiometabolic disease, obesity, depression, and immune dysfunction[6, 7]. Ambient light is the dominant circadian zeitgeber, entraining the suprachiasmatic nucleus and modulating melatonin secretion[8, 9]. Wrist-worn photosensors enable continuous, objective characterization of personal light exposure in free-living conditions across multiple days, capturing patterns that correlate with sleep timing, social jet lag, and health outcomes[10–13]. Behavioral and circadian pathways connecting tobacco smoke exposure to ambient light patterns are plausible. Nicotine has been shown to modulate circadian clock gene expression in animal models[14]. Current smokers report later chronotypes, shorter sleep duration, and greater sleep fragmentation in observational studies[15–17]. These associations suggest that tobacco smoke co-exposure may co-occur with behavioral patterns, including altered activity timing and indoor light environments,that affect personal light–dark cycle exposure. However, this relationship has not been examined using objective biomarker exposure and objective wrist-worn light measurement in a nationally representative sample.

Prior studies have examined smoking in relation to sleep duration, chronotype, and circadian disruption,[15–18] and separately, wrist-worn light data have been used to characterize circadian-relevant personal light exposure[10–12]. However, prior nationally representative work has not combined serum cotinine with NHANES wrist-worn light measures to examine free-living light–dark exposure patterns. The key contribution of the present study is linking an objective biomarker of tobacco smoke exposure to objectively measured, multi-day personal light environment in a complex-survey-weighted nationally representative sample, a combination not previously reported. We hypothesized that higher cotinine would co-occur with disrupted light patterns, specifically higher nighttime light and lower evening light,after sociodemographic adjustment, consistent with cotinine marking a broader behavioral-exposure phenotype.

## 2. Methods

### 2.1. Study Design and Population

This cross-sectional study used NHANES 2011–2012 and 2013–2014 (cycles G and H), conducted by the National Center for Health Statistics (NCHS) using complex multi-stage probability sampling[19].

Inclusion criteria were: adults aged ≥20 years with (1) valid serum cotinine; (2) valid wrist-worn PAM light data (≥4 valid monitoring days, ≥40 valid minutes per hour); (3) non-missing survey design variables (SDMVSTRA, SDMVPSU, WTMEC2YR); and (4) complete age, sex, and race/ethnicity data. Pregnant participants were excluded. Final analytic *N* = 6,937.

### 2.2. Primary Exposure: Serum Cotinine

Serum cotinine was measured by HPLC–APCI MS/MS (LLOD = 0.011 ng/mL)[3, 20]. For regression analyses, cotinine was log_2_-transformed after adding 1, i.e., log_2_(cotinine + 1), to address right skew and near-zero values. For descriptive analyses, three categories were defined based on established biomarker thresholds:[3, 4] (1) *Low/No Exposure* (<0.05 ng/mL); (2) *cotinine 0*.*05–* <*3 ng/mL* (labeled possible secondhand smoke exposure); and (3) *cotinine* ≥*3 ng/mL* (labeled likely active smoking). These labels reflect probabilistic population-level patterns; cotinine alone cannot definitively identify individual exposure source.

### 2.3. Ambient Light Outcomes

The NHANES PAM substudy used an Actigraph GT3X+ on the non-dominant wrist for up to 7 days, recording ambient illuminance (lux) in one-hour epochs[21]. Clock hours were derived from the device start time (PAXFTIME field in the NHANES accelerometry header) combined with a global sequential hour index within each participant. A valid hour required ≥40 valid wear minutes (PAXVMH). Participants needed ≥4 valid days with ≥16 valid hours each. Hours not meeting the valid-wear criterion were excluded before computing light summaries; participant-level outcomes were calculated as the mean of valid hours and valid days only.

Seven primary light outcomes were derived (light values transformed as natural logarithm of lux+1, i.e., ln(lux+1), to normalize right-skewed distributions):

1. Mean nighttime light [ln(lux + 1), 00:00–05:59]
2. Mean evening light [ln(lux + 1), 20:00–23:59]
3. Mean morning light [ln(lux + 1), 06:00–09:59]
4. Day–night contrast (06:00–19:59 minus nighttime mean)
5. Lux-weighted light centroid timing (hours, 0–23)
6. SD of daily centroid timing (hours)
7. Inter-day light regularity (Pearson *r* of consecutive day profiles)

### 2.4. Covariates

**Model 1:** age (continuous), sex (male/female), race/ethnicity (5 categories). **Model 2 (primary)**: Model 1 plus education (3 levels), poverty-income ratio (PIR, continuous), BMI (continuous), and survey cycle. **Model 3a (Exploratory, no alcohol)**: Model 2 plus sleep duration (hours), log-caffeine (mg/day), and PHQ-9 depression score. **Model 3b (Exploratory, full behavioral)**: Model 3a plus alcohol consumption (drinks/day). Alcohol data were missing for 37.6% of participants, reducing the analytic sample from *N* ≈ 6,350 to *N* ≈ 3,866. Presenting Models 3a and 3b separately allows readers to distinguish covariate-adjustment effects from sample-restriction effects. Sleep duration and PHQ-9 may lie downstream of cotinine/smoking and should be considered possible behavioral mediators rather than pure confounders; both Model 3 specifications are therefore labeled exploratory. **Model 4**: Model 3b plus employment status (employed/not employed).

### 2.5. Statistical Analysis

All analyses used four-year pooled MEC weights (WTMEC4YR = WTMEC2YR / 2), stratification by SDMVSTRA, and clustering by SDMVPSU per NCHS guidelines[19, 22]. Lonely PSU strata were handled using the “adjust” (centered) option; survey degrees of freedom = 32. Survey-weighted linear regression used the svyglm function from the R survey package[23]. FDR correction used the Benjamini–Hochberg procedure across the 7 primary outcomes within each model (*p*_FDR_ < 0.05 threshold)[24]. Model 2 is the primary specification.

Sensitivity analyses included: (1) adding self-reported smoking status to Model 2; (2) stratification by smoking status; (3) cotinine as categorical (three groups vs. low/no); (4) Model 2 restricted to the Model 3 complete-case sample; (5) winsorizing nighttime light at the 99th percentile; and (6) exploratory sex-stratified analyses with formal interaction testing. Interaction tests were not pre-specified and are labeled exploratory; no correction for multiple interaction tests was applied.

## 3. Results

### 3.1. Sample Characteristics

The analytic sample included 6,937 adults (survey-weighted mean age 52.4 years; 38.4% female). Unweighted counts by cotinine category: Low (<0.05 ng/mL) *N* = 3,830; Intermediate (0.05–<3 ng/mL) *N* = 1,259; High (≥3 ng/mL) *N* = 1,848. Participants with higher cotinine were more likely to be male, less educated, with lower poverty-income ratios and higher PHQ-9 scores. Mean nighttime light increased and mean evening light decreased across cotinine categories (Table 1).

**Table 1:** Weighted means (±SD) or weighted percentages by serum cotinine category, NHANES 2011–2014. Column *N* values are unweighted; all statistics are survey-weighted.

| Characteristic | Low cotinine<br>($<0.05$ ng/mL)<br>$N = 3,830$ | Intermediate cotinine<br>($0.05$–$<3$ ng/mL)<br>$N = 1,259$ | High cotinine<br>($\geq 3$ ng/mL)<br>$N = 1,848$ |
| --- | --- | --- | --- |
| Age, years | $55.5 \pm 15.5$ | $49.4 \pm 17.6$ | $47.2 \pm 14.5$ |
| Female sex, % | 44.8 | 33.7 | 26.9 |
| Non-Hispanic White, % | 73.2 | 60.9 | 69.0 |
| Non-Hispanic Black, % | 7.0 | 15.6 | 13.6 |
| Mexican American, % | 7.6 | 8.5 | 6.2 |
| Other Hispanic, % | 5.6 | 6.1 | 4.7 |
| College+ education, % | 40.8 | 21.7 | 13.2 |
| Poverty-income ratio | $3.40 \pm 1.57$ | $2.66 \pm 1.63$ | $2.37 \pm 1.58$ |
| BMI, kg/m<sup>2</sup> | $29.2 \pm 6.3$ | $30.6 \pm 7.5$ | $28.4 \pm 6.5$ |
| Sleep duration, h | $7.0 \pm 1.2$ | $7.0 \pm 1.3$ | $6.7 \pm 1.5$ |
| PHQ-9 score | $2.5 \pm 3.8$ | $3.0 \pm 4.3$ | $3.9 \pm 4.9$ |
| Nighttime light, $\ln(\text{lux} + 1)$ | $0.84 \pm 1.02$ | $0.99 \pm 1.03$ | $1.10 \pm 1.09$ |
| Evening light, $\ln(\text{lux} + 1)$ | $3.88 \pm 1.75$ | $3.42 \pm 1.73$ | $3.37 \pm 1.72$ |
| Day–night contrast | $6.55 \pm 1.99$ | $6.13 \pm 2.10$ | $6.13 \pm 2.26$ |

BMI = body mass index; PIR = poverty-income ratio.
Intermediate cotinine ( $0.05$ – $<3$ ng/mL) and high cotinine ( $\geq 3$ ng/mL) categories reflect approximate population-level thresholds for possible secondhand smoke exposure and likely active smoking, respectively. Cotinine concentration alone cannot definitively identify individual exposure source; these labels are descriptive.

### 3.2. Primary Continuous Cotinine Associations (Models 1–4)

Table 2 presents survey-weighted regression results. In Model 2 (primary, *N* = 6,350), two outcomes were FDR-significant: nighttime light (*β* = +0.024, 95% CI: 0.010, 0.038; *p*_FDR_ = 0.014) and evening light (*β* = −0.031, 95% CI: −0.055, −0.008; *p*_FDR_ = 0.042).Morning light was significant in Model 1 (*p*_FDR_ = 0.002) but not after fuller socioeconomic adjustment in Model 2 (*p*_FDR_ = 0.216). No other outcomes were FDR-significant in Model 2.

**Table 2:**
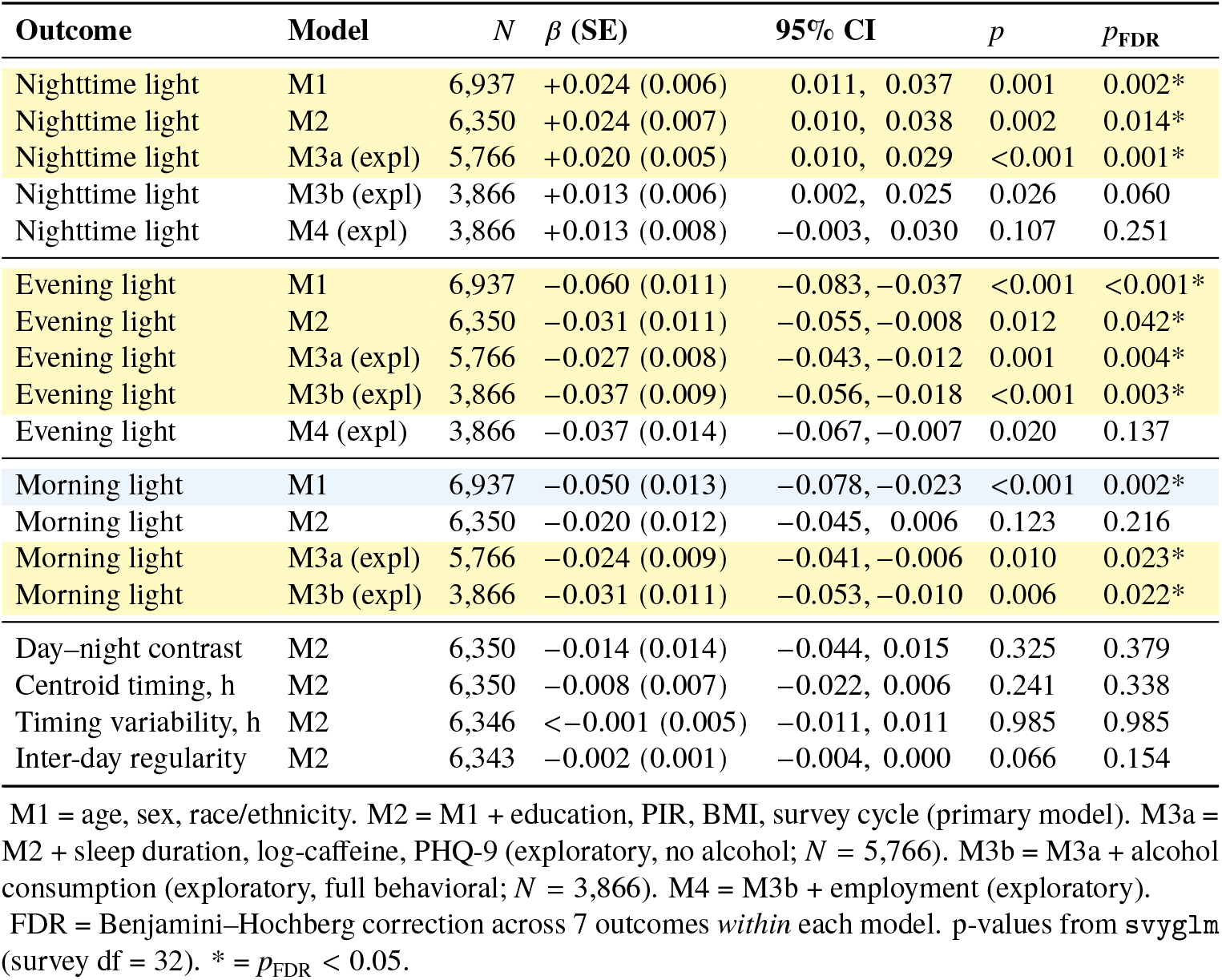
Survey-weighted regression coefficients for log_2_(cotinine + 1) predicting ambient light outcomes across models, NHANES 2011–2014. Yellow = FDR-significant (*p*_FDR_ < 0.05). Blue = FDR-significant in Model 1 only. M3a and M3b are exploratory; M3b adds alcohol to M3a, reducing sample by 33%.

| Outcome | Model | N | $\beta$ (SE) | 95% CI | p | $p_{\text{FDR}}$ |
| --- | --- | --- | --- | --- | --- | --- |
| Nighttime light | M1 | 6,937 | +0.024 (0.006) | 0.011, 0.037 | 0.001 | 0.002* |
| Nighttime light | M2 | 6,350 | +0.024 (0.007) | 0.010, 0.038 | 0.002 | 0.014* |
| Nighttime light | M3a (expl) | 5,766 | +0.020 (0.005) | 0.010, 0.029 | <0.001 | 0.001* |
| Nighttime light | M3b (expl) | 3,866 | +0.013 (0.006) | 0.002, 0.025 | 0.026 | 0.060 |
| Nighttime light | M4 (expl) | 3,866 | +0.013 (0.008) | −0.003, 0.030 | 0.107 | 0.251 |
| Evening light | M1 | 6,937 | −0.060 (0.011) | −0.083, −0.037 | <0.001 | <0.001* |
| Evening light | M2 | 6,350 | −0.031 (0.011) | −0.055, −0.008 | 0.012 | 0.042* |
| Evening light | M3a (expl) | 5,766 | −0.027 (0.008) | −0.043, −0.012 | 0.001 | 0.004* |
| Evening light | M3b (expl) | 3,866 | −0.037 (0.009) | −0.056, −0.018 | <0.001 | 0.003* |
| Evening light | M4 (expl) | 3,866 | −0.037 (0.014) | −0.067, −0.007 | 0.020 | 0.137 |
| Morning light | M1 | 6,937 | −0.050 (0.013) | −0.078, −0.023 | <0.001 | 0.002* |
| Morning light | M2 | 6,350 | −0.020 (0.012) | −0.045, 0.006 | 0.123 | 0.216 |
| Morning light | M3a (expl) | 5,766 | −0.024 (0.009) | −0.041, −0.006 | 0.010 | 0.023* |
| Morning light | M3b (expl) | 3,866 | −0.031 (0.011) | −0.053, −0.010 | 0.006 | 0.022* |
| Day–night contrast | M2 | 6,350 | −0.014 (0.014) | −0.044, 0.015 | 0.325 | 0.379 |
| Centroid timing, h | M2 | 6,350 | −0.008 (0.007) | −0.022, 0.006 | 0.241 | 0.338 |
| Timing variability, h | M2 | 6,346 | <−0.001 (0.005) | −0.011, 0.011 | 0.985 | 0.985 |
| Inter-day regularity | M2 | 6,343 | −0.002 (0.001) | −0.004, 0.000 | 0.066 | 0.154 |

M1 = age, sex, race/ethnicity. M2 = M1 + education, PIR, BMI, survey cycle (primary model). M3a = M2 + sleep duration, log-caffeine, PHQ-9 (exploratory, no alcohol; $N = 5,766$ ). M3b = M3a + alcohol consumption (exploratory, full behavioral; $N = 3,866$ ). M4 = M3b + employment (exploratory).
FDR = Benjamini–Hochberg correction across 7 outcomes *within* each model. p-values from svyglm (survey df = 32). \* = $p_{\text{FDR}} < 0.05$ .

To interpret effect magnitude: a *β* = +0.024 on natural-log-scale ln(lux + 1) per log_2_(cotinine + 1) unit corresponds to approximately a 2.4% higher nighttime lux per log_2_-unit cotinine increase, after back-transformation [*e*^0.024^ − 1 ≈ 0.024]. While individually modest, this association operates across a high-prevalence exposure (tobacco smoke) and a ubiquitous environmental factor (artificial light), suggesting potential population-level relevance. The association marks a behavioral co-exposure pattern rather than a direct biological effect of cotinine on light environment.

To disentangle the effect of behavioral covariate adjustment from alcohol-driven sample restriction, we report two exploratory models. Model 3a added sleep duration, log-caffeine, and PHQ-9 without alcohol (*N* = 5,766): nighttime light (*β* = +0.020, 95% CI: 0.010, 0.029; *p*_FDR_ = 0.001) and evening light (*β* = −0.027, 95% CI: −0.043, −0.012; *p*_FDR_ = 0.004) remained FDR-significant. Morning light also became FDR-significant in Model 3a (*β* = −0.024; *p*_FDR_ = 0.023), reflecting adjustment for sleep duration. Model 3b further added alcohol (*N* = 3,866, a 33% sample reduction due to 37.6% missing alcohol data): nighttime light attenuated to *β* = +0.013 and lost FDR significance (*p*_FDR_ = 0.060), while evening light remained FDR-significant (*β* = −0.037; *p*_FDR_ = 0.003) and morning light also remained significant (*β* = −0.031; *p*_FDR_ = 0.022). The nighttime light attenuation from M2 to M3b therefore reflects both genuine behavioral covariate adjustment and sample restriction from missing alcohol data; these effects cannot be fully separated with complete-case analysis alone.

### 3.3. Categorical Exposure-Gradient Analysis

Modeling cotinine as categories revealed a categorical exposure-gradient pattern for nighttime light and a threshold pattern for evening and morning light (Table 3). Night-time light showed progressively higher values: +0.096 log(lux+1) for the 0.05–<3 ng/mL group vs. Low/No (*p* = 0.018), and +0.209 for the ≥3 ng/mL group vs. Low/No (*p* < 0.001). For evening and morning light, both exposed groups were significantly lower than Low/No, but the two exposed groups were similar to each other (evening: −0.274 vs. −0.281), indicating a threshold rather than a linear gradient.

**Table 3:** Categorical cotinine associations with ambient light outcomes (reference = Low/No <0.05 ng/mL), Model 2 adjustment, NHANES 2011–2014. *N* = 6,350 for all models.

| Outcome | Cotinine group vs Low (<0.05 ng/mL) | $\beta$ | CI low | CI high | $p$ |
| --- | --- | --- | --- | --- | --- |
| Nighttime | Intermediate (0.05–<3) | +0.096 | +0.018 | +0.173 | 0.018 |
| Nighttime | High ($\geq 3$ ng/mL) | +0.209 | +0.141 | +0.277 | <0.001 |
| Evening | Intermediate (0.05–<3) | –0.274 | –0.403 | –0.146 | <0.001 |
| Evening | High ($\geq 3$ ng/mL) | –0.281 | –0.393 | –0.168 | <0.001 |
| Morning | Intermediate (0.05–<3) | –0.218 | –0.366 | –0.071 | 0.005 |
| Morning | High ($\geq 3$ ng/mL) | –0.258 | –0.387 | –0.129 | <0.001 |

Model 2 adjustment: age, sex, race/ethnicity, education, PIR, BMI, survey cycle. Cotinine category thresholds are approximate; they do not definitively classify exposure source.

### 3.4. Primary Result Figures

Figure 1 shows the Model 2 forest plot across all seven outcomes. Figure 2 shows survey-weighted predicted nighttime light by cotinine category.

**Figure 1.**
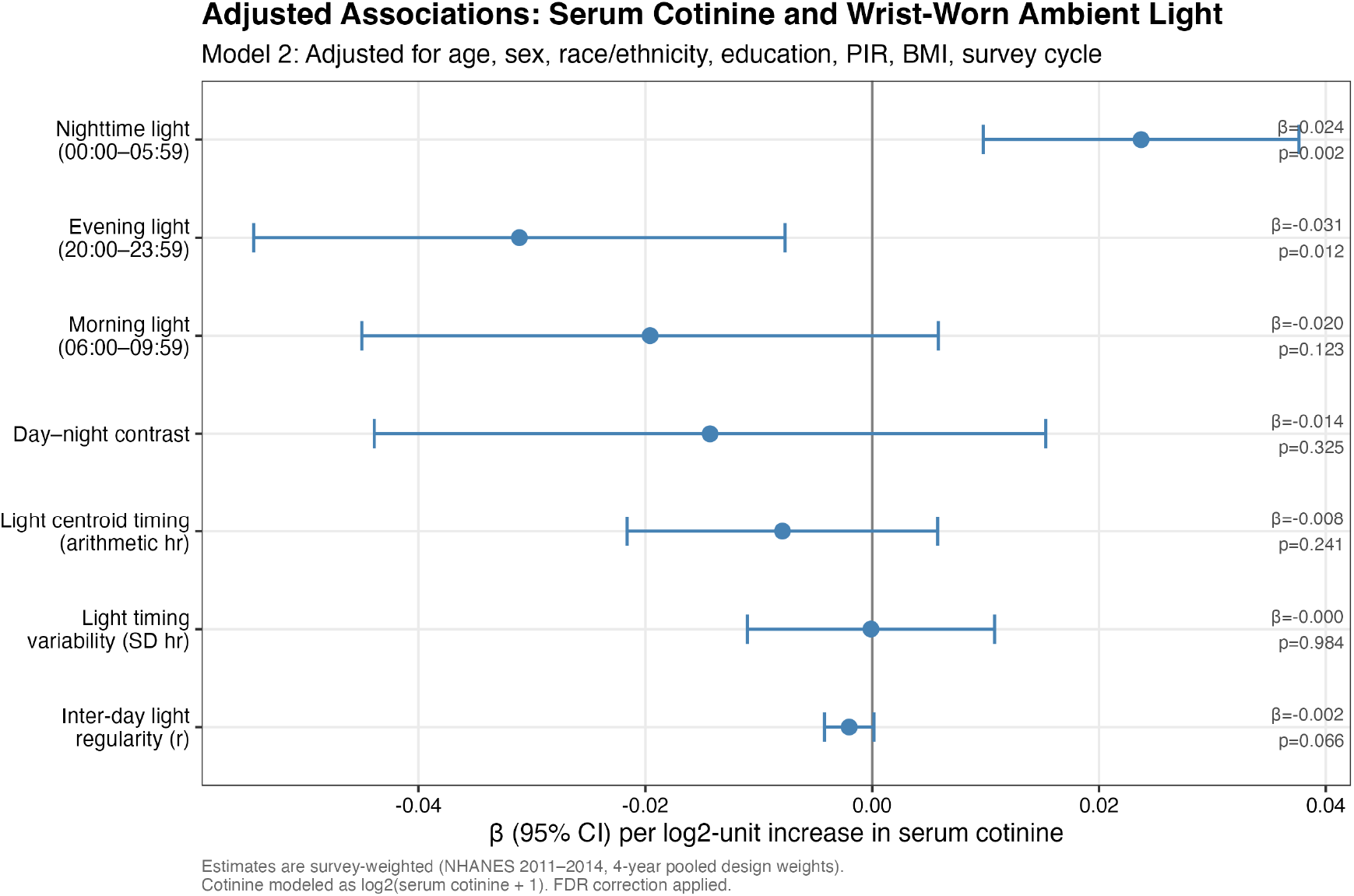
Forest plot: survey-weighted *β* coefficients (95% CI) for log_2_(cotinine + 1) and all 7 ambient light outcomes, Model 2 (*N* = 6,350), NHANES 2011–2014. Asterisks (*) denote FDR-significant outcomes (*p*_FDR_ < 0.05): nighttime light and evening light. All outcomes are shown to reduce selective-reporting concerns.

**Figure 2.**
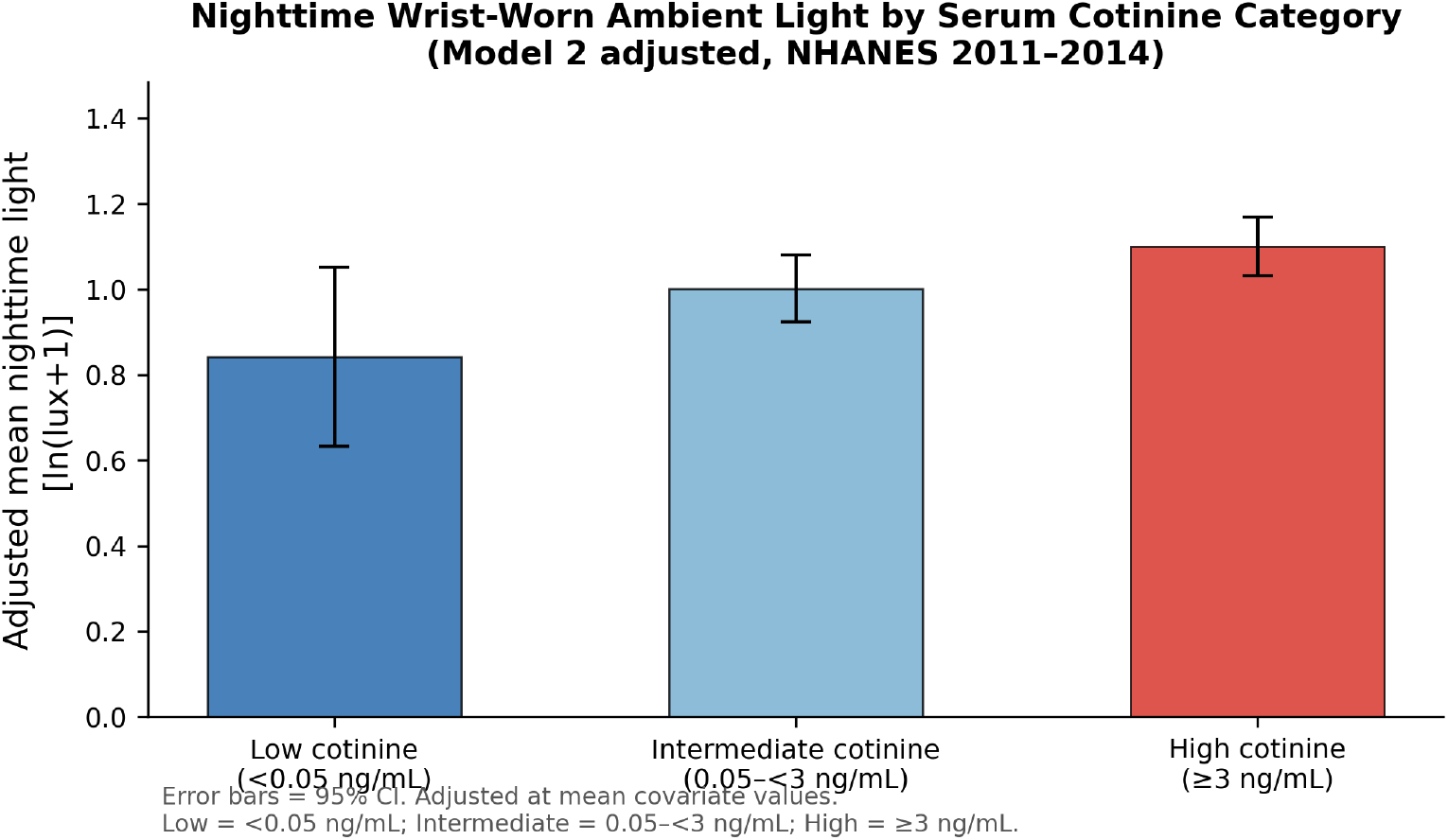
Adjusted mean nighttime wrist-worn ambient light [ln(lux + 1)] by serum cotinine category, Model 2 covariates. Error bars = 95% CIs. Low = <0.05 ng/mL; Intermediate = 0.05–<3 ng/mL (approximates possible secondhand smoke exposure at the population level); High = ≥3 ng/mL (approximates likely active smoking at the population level). NHANES 2011–2014.

### 3.5. Sensitivity Analyses

Table 4 summarizes key sensitivity analyses. Results were robust to winsorizing nighttime light at the 99th percentile (*β* = +0.023, *p* = 0.002). When self-reported smoking status was added to Model 2, the cotinine–nighttime association attenuated substantially (*β* = +0.001, *p* = 0.94, *N* = 6,344), consistent with the high collinearity between serum cotinine and smoking status. This indicates that the continuous cotinine association captures largely the same information as categorical smoking status when both are included simultaneously, and that the cotinine–nighttime association in Model 2 is not independent of smoking behavior. The evening light association persisted after smoking-status adjustment (*β* = −0.043, *p* = 0.025). Among never-smokers only (*N* = 3,345), the evening light association remained nominally significant (*β* = −0.041, *p* = 0.034).

**Table 4:**
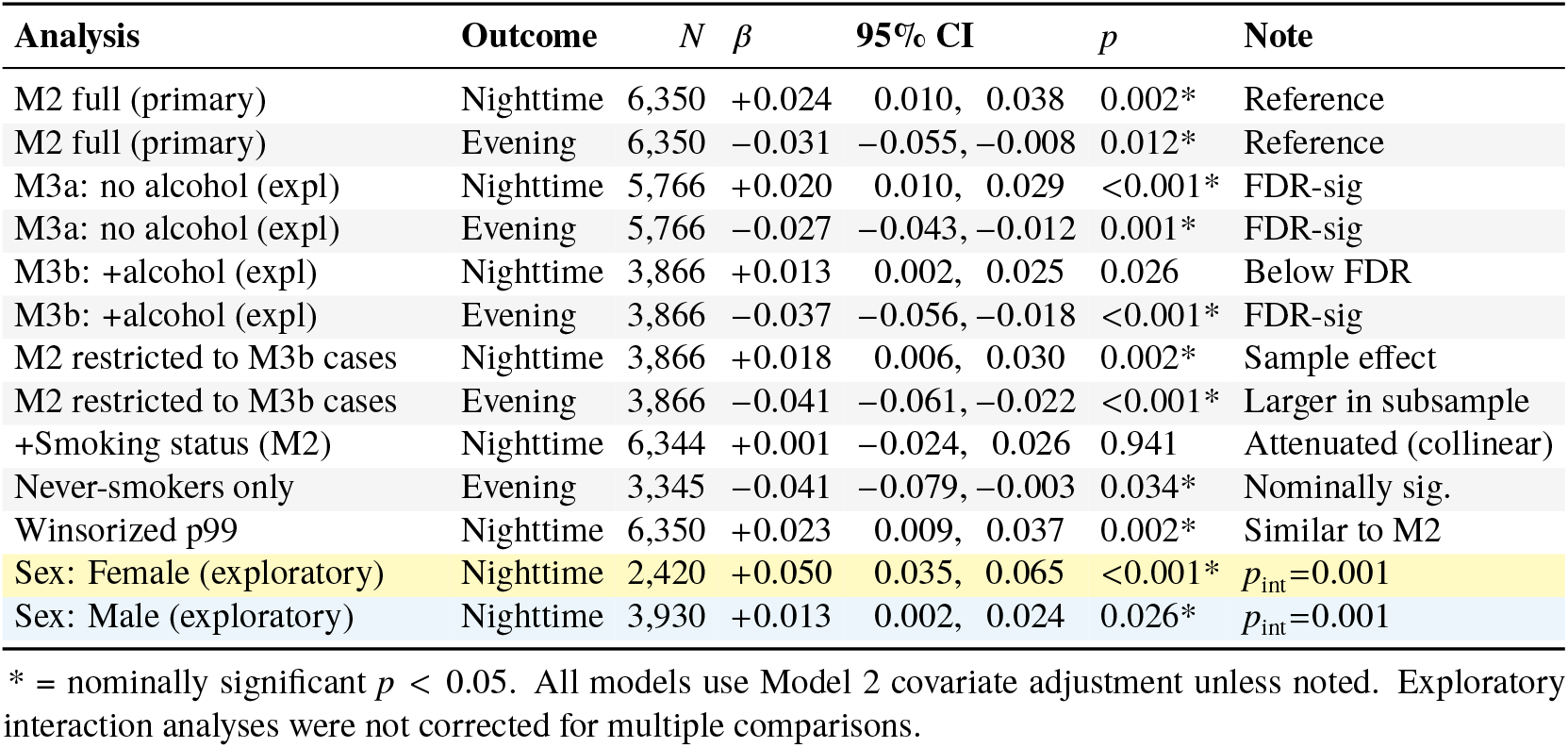
Sensitivity analysis summary for nighttime and evening light (primary FDR-significant outcomes), NHANES 2011–2014.

| Analysis | Outcome | $N$ | $\beta$ | 95% CI | $p$ | Note |
| --- | --- | --- | --- | --- | --- | --- |
| M2 full (primary) | Nighttime | 6,350 | +0.024 | 0.010, 0.038 | 0.002* | Reference |
| M2 full (primary) | Evening | 6,350 | –0.031 | –0.055, –0.008 | 0.012* | Reference |
| M3a: no alcohol (expl) | Nighttime | 5,766 | +0.020 | 0.010, 0.029 | <0.001* | FDR-sig |
| M3a: no alcohol (expl) | Evening | 5,766 | –0.027 | –0.043, –0.012 | 0.001* | FDR-sig |
| M3b: +alcohol (expl) | Nighttime | 3,866 | +0.013 | 0.002, 0.025 | 0.026 | Below FDR |
| M3b: +alcohol (expl) | Evening | 3,866 | –0.037 | –0.056, –0.018 | <0.001* | FDR-sig |
| M2 restricted to M3b cases | Nighttime | 3,866 | +0.018 | 0.006, 0.030 | 0.002* | Sample effect |
| M2 restricted to M3b cases | Evening | 3,866 | –0.041 | –0.061, –0.022 | <0.001* | Larger in subsample |
| +Smoking status (M2) | Nighttime | 6,344 | +0.001 | –0.024, 0.026 | 0.941 | Attenuated (collinear) |
| Never-smokers only | Evening | 3,345 | –0.041 | –0.079, –0.003 | 0.034* | Nominally sig. |
| Winsorized p99 | Nighttime | 6,350 | +0.023 | 0.009, 0.037 | 0.002* | Similar to M2 |
| Sex: Female (exploratory) | Nighttime | 2,420 | +0.050 | 0.035, 0.065 | <0.001* | $p_{\text{int}}=0.001$ |
| Sex: Male (exploratory) | Nighttime | 3,930 | +0.013 | 0.002, 0.024 | 0.026* | $p_{\text{int}}=0.001$ |

\* = nominally significant $p < 0.05$ . All models use Model 2 covariate adjustment unless noted. Exploratory interaction analyses were not corrected for multiple comparisons.

Exploratory sex-stratified analyses identified a significant cotinine-by-sex interaction for nighttime light (*p*_interaction_ = 0.001). The association was stronger in females (*β* = +0.050, 95% CI: 0.035, 0.065) than males (*β* = +0.013, 95% CI: 0.002, 0.024), while the evening light association was significant in males (*β* = −0.037, *p* < 0.001) but not females (*β* = −0.008, *p* = 0.531). Because these interaction tests were not pre-specified and were not corrected for multiple comparisons, they are hypothesis-generating and require replication in independent samples.

## 4. Discussion

In this nationally representative cross-sectional analysis of 6,937 U.S. adults, higher serum cotinine was associated with higher nighttime ambient light and lower evening ambient light after sociodemographic adjustment (Model 2, FDR-corrected). The most parsimonious interpretation is that cotinine marks a behavioral-exposure phenotype characterized by later nocturnal activity, greater artificial light at night, and reduced evening outdoor exposure,rather than a direct pharmacological effect of nicotine on light environment. A categorical exposure-gradient pattern across cotinine groups, persistence of the evening light association in never-smokers, and a hypothesis-generating sex interaction collectively support the behavioral and circadian plausibility of these associations.

Higher nighttime light with greater cotinine likely reflects behavioral patterns co-occurring with tobacco use, including later nocturnal activity and greater exposure to artificial light at night[18, 25]. Lower evening light with higher cotinine could reflect less time in well-lit social or outdoor environments during the evening, reducing the photoentraining signal that normally anchors the circadian pacemaker[26].

### 4.1. Attenuation in Exploratory Models: Confounding, Mediation, or Selection?

The M3a/M3b decomposition provides key mechanistic insight. Nighttime light remained FDR-significant in Model 3a (*N* = 5,766, no alcohol), indicating that sleep duration, caffeine, and PHQ-9 adjustment alone does not eliminate the association. Nighttime light lost FDR significance only in Model 3b, which added alcohol and reduced the sample to *N* = 3,866 due to 37.6% missing alcohol data. This suggests the nighttime light attenuation reflects a combination of: (1) partial confounding or mediation by alcohol use, and (2) sample-selection bias from complete-case restriction to participants with alcohol data, who are younger, higher-cotinine, and higher-income than the full sample. In contrast, the evening light association was robust across Models 3a and 3b, remaining FDR-significant in both (*p*_FDR_ = 0.004 and *p*_FDR_ = 0.003 respectively), suggesting it is less sensitive to either behavioral confounders or the alcohol-driven sample restriction. Sleep duration and PHQ-9 may lie down-stream of cotinine/smoking and could act as partial mediators; Model 3a and 3b therefore represent exploratory behavioral-adjustment bounds rather than definitive causal estimates. Model 2 remains the primary specification.

### 4.2. Sex Differences (Exploratory)

The significant sex interaction for nighttime light (*p*_interaction_ = 0.001) is hypothesis-generating. The stronger cotinine–nighttime association in females (*β* = +0.050) may reflect sex-specific behavioral patterns associated with tobacco use, or differences in indoor versus outdoor activity timing. These patterns should be treated with caution given the post-hoc nature of the analysis and the absence of multiple-testing correction across interaction tests.

### 4.3. Categorical Patterns

The categorical analysis showed that even participants with cotinine 0.05–<3 ng/mL had significantly higher nighttime light and lower evening and morning light compared with the Low/No group, suggesting the association is not limited to heavy smokers. However, the evening and morning light pattern was a threshold effect (both exposed groups were similarly lower than Low/No) rather than a linear gradient, which tempers the dose-response interpretation for those outcomes.

### 4.4. Limitations

The cross-sectional design precludes causal inference; reverse causation cannot be excluded. Wrist-worn lux reflects ambient wrist illuminance rather than retinal irradiance driving circadian photoentrainment,[27] and may be attenuated by wrist position, clothing, and indoor–outdoor context; field comparisons suggest moderate but imperfect correspondence between wrist-worn and head-level light measures[13]. The primary driver of Model 3 sample reduction is missing alcohol data (37.6%), introducing selection bias. Self-reported covariates are subject to measurement error. Cotinine category thresholds are approximate, and the 0.05–<3 ng/mL range encompasses diverse exposure sources including e-cigarette use and nicotine replacement therapy[28].Shift work is among the most important unmeasured confounders: shift workers have substantially elevated nighttime light exposure by occupational necessity and have higher smoking prevalence than day workers. We did not have harmonized shift-work status available for this analysis, meaning that the nighttime light association with cotinine could partially or wholly reflect unmeasured occupational schedules rather than tobacco-related behavioral patterns. We also lacked data on housing conditions, neighborhood light environment, nighttime caregiving responsibilities, and indoor smoking restrictions, all of which could confound or modify the observed associations.

Multiple imputation for missing alcohol data was not performed and represents an important robustness check for future work.

## 5. Conclusions

Higher serum cotinine concentrations were associated with higher nighttime and lower evening wrist-worn ambient light in a nationally representative U.S. adult sample, after sociodemographic adjustment. Attenuation after behavioral adjustment suggests possible partial mediation, sample selection effects, or both. A categorical exposure-gradient pattern for nighttime light and persistence of the evening light association in never-smokers support the behavioral and circadian relevance of these findings. The cross-sectional design and potential for residual confounding preclude causal inference. Longitudinal studies with complete behavioral covariate data and formal mediation analyses are needed to clarify the temporal ordering and mechanisms linking tobacco smoke exposure, smoking-related behaviors, and personal light–dark cycle patterns.

## Data Availability Statement

All NHANES data used in this analysis are publicly available from the National Center for Health Statistics at https://wwwn.cdc.gov/nchs/nhanes/. Analysis code will be made publicly available on GitHub upon publication.

## Code Availability

Analysis code used to construct light outcomes from NHANES PAM accelerometry files and to reproduce all regression models, tables, and figures will be deposited in a public GitHub repository at the time of publication. Code is available from the corresponding author upon reasonable request prior to that time.

## Ethics Statement

NHANES protocols were approved by the National Center for Health Statistics Research Ethics Review Board. All participants provided written informed consent. This secondary analysis of publicly available, de-identified data did not require additional IRB review.

## Author Contributions

A.C.W.: conceptualization, data curation, formal analysis, writing, original draft, writing, review and editing. C.L., A.P., L.Y., Y.C.: writing, review and editing. All authors reviewed and approved the final manuscript.

